# Treatment of sarcoidosis with cutaneous involvement with tofacitinib

**DOI:** 10.1101/2021.07.01.21259700

**Authors:** William Damsky, Alice Wang, Bryan D. Young, Ruveyda Ayasun, Changwan Ryu, Meaghan K. McGeary, Ramesh Fazzone-Chettiar, Darko Pucar, Mridu Gulati, Edward J. Miller, Marcus Bosenberg, Richard A. Flavell, Brett King

## Abstract

Sarcoidosis is an idiopathic inflammatory disorder that is commonly treated with glucocorticoids and there are no approved steroid-sparing medications. There is emerging evidence that Janus kinase (JAK) inhibitors, which inhibit JAK-dependent cytokine activity, may hold promise in sarcoidosis. In this open-label trial, 10 patients with recalcitrant sarcoidosis with cutaneous involvement were treated with tofacitinib 5 mg twice daily. There was no washout period and patients were permitted to continue, taper, or discontinue other treatments. The primary outcome was the change in the Cutaneous Sarcoidosis Activity and Morphology Instrument (CSAMI) activity score after 6 months. Change in internal organ disease activity was also assessed using total lesion glycolysis (TLG) determined by full-body positron emission tomography. A mean reduction in the CSAMI activity score of 82.7% was observed, with 6 patients showing a complete response. Internal organ response data was available in 8 patients; a decrease in TLG of ≥50% was noted in 5 patients, with complete or near complete resolution in 3 (>98% reduction in TLG). Patients were generally able to significantly taper or discontinue their baseline immunosuppressive regimen, which included prednisone in 5 patients. Single cell RNA-sequencing, bulk RNA-sequencing, and high-throughput proteomic analyses were performed on skin and blood as a function of treatment in order to delineate changes in immunologic signals with therapy. We identified CD4+ T cell derived IFN-γ as a central cytokine driver of sarcoidosis and inhibition of its activity was achieved with tofacitinib and correlated closely with clinical improvement. Tofacitinib appears to have impressive activity in treatment of sarcoidosis and likely acts by inhibiting IFN-γ, larger, controlled studies are warranted.

## Introduction

Sarcoidosis is an inflammatory disorder that most commonly affects the lungs; however, any organ including the skin can be involved. Glucocorticoid-based regimens (prednisone and corticotropin gel) are the only approved therapies. Prednisone remains the recommended first-line treatment for both pulmonary and extra-thoracic sarcoidosis^1^. However, this is not ideal as chronic therapy is often required and glucocorticoid-associated toxicities are common^2^. Methotrexate is commonly used as a steroid-sparing agent, but it is often inadequate^1^. TNF-α inhibitors have also been evaluated; however, the benefit in controlled trials has been marginal, with several studies finding no benefit^3-8^. TNF-α inhibitors can also induce or exacerbate sarcoidosis^9^.

A hallmark of sarcoidosis is the non-caseating granuloma, which is observed microscopically in affected tissues. Although the pathogenesis of granuloma formation in sarcoidosis is complex, it appears to involve coordinated activity of several cytokines, chemokines, and other signals ^10^. Of these, many cytokines including IFN-γ, Type I IFNs (IFN-α/β), IL-2, IL-4, IL-6, IL-12, IL-13, IL-23 and GM-CSF signal via the JAK – signal transducer and activator of transcription (STAT) pathway. Indeed, JAK-STAT pathway activation has been reported in tissues and blood of patients with sarcoidosis^11-15^. JAK inhibitors are oral small molecules that attenuate the activity of JAK-STAT-dependent cytokines. We and others have recently described effective treatment of individual patients with sarcoidosis using tofacitinib (JAK1/2/3 inhibitor) or ruxolitinib (JAK1/2 inhibitor)^14-20^.

We report 10 patients with long-standing, recalcitrant sarcoidosis with cutaneous involvement treated with tofacitinib for 6 months. In all 10 patients, disease control with a tofacitinib-based regimen was superior to the preceding immunotherapeutic regimen, particularly for skin involvement. Molecular analyses were performed and identified IFN-γ as a key driver of sarcoidosis pathogenesis and central target of tofacitinib. Overall, JAK inhibition appears to be effective in sarcoidosis and larger studies appear to be warranted.

## Methods

### Population and Study Design

This is an open-label trial conducted at a Yale University. Patients were at least 18 years old and had a diagnosis of sarcoidosis with a supportive skin biopsy and a CSAMI activity score^21^ ≥10. Nine patients also had internal organ involvement **(Table 1)**. Any preceding treatment-regimen for sarcoidosis had to be stable over the preceding 3 months. All patients provided written informed consent. Full details are available in the **Supplementary Appendix**.

**Table 1.**
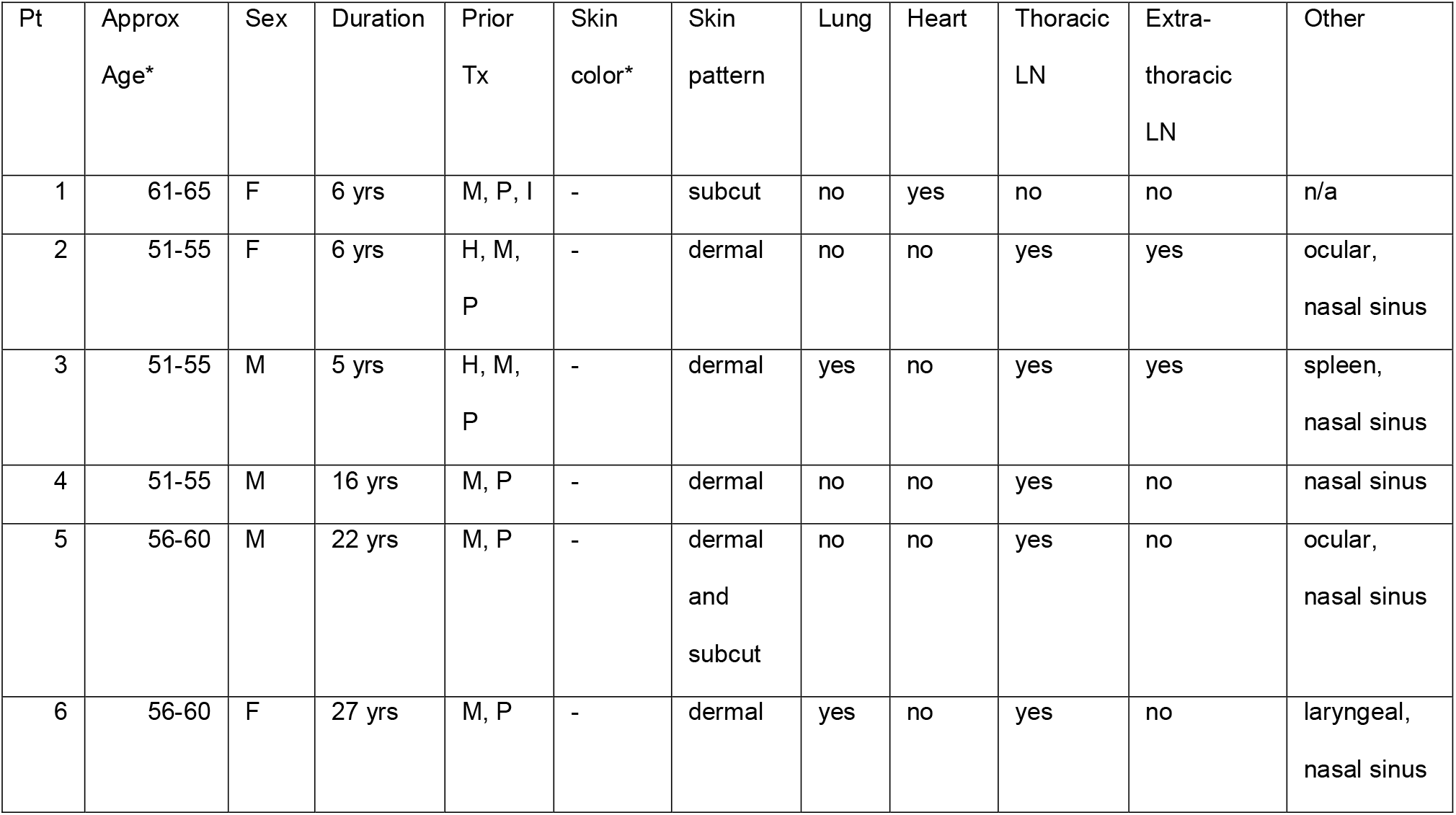
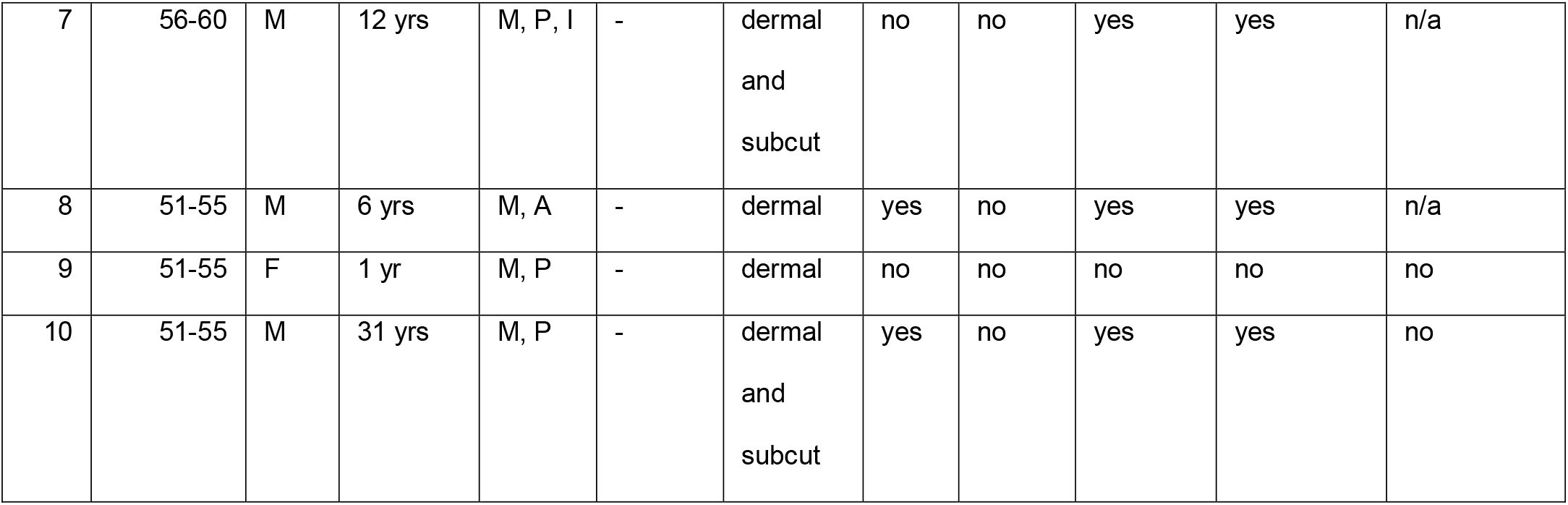
Patient demographics and characteristics. Prior treatments (tx) encompass all previously utilized therapies. The therapeutic regimen for each patient at the beginning of the study is summarized in Figure 1. Skin color depicted by Fitzpatrick phototype (I-VI). H: hydroxychloroquine, P: prednisone, M: methotrexate, I: infliximab, A: azathioprine. *Some potentially identifying patient information has been removed.

### Study treatment and endpoints

There was no washout period and baseline data were obtained while taking the preceding immunosuppressive regimen **(Table 1)**. Tofacitinib 5 mg twice daily was initiated, and patients were evaluated after 1, 3, and 6 months of treatment. Tapering or discontinuation of other sarcoidosis treatment(s) was permitted. The primary endpoint was change in the CSAMI activity score after 6 months. Change in internal organ involvement was determined by whole body PET – computed tomography (CT)^16^. Total lesion glycolysis (TLG) was determined in using MIM software^22^. Change in skin-related quality of life (Skindex-16)^23^ was also assessed. Molecular parameters were assessed at baseline and after 6 months of treatment. Full details are described in the **Supplementary Appendix**.

### Study Oversight

The study was approved by the Yale Institutional Review Board and performed in accordance with the protocol and with the principles of the Declaration of Helsinki and local regulations. All authors had access to the primary data and approved the decision to submit the manuscript.

## Results

### Tofacitinib leads to improvement in cutaneous sarcoidosis

All 10 patients had cutaneous sarcoidosis. The average age was 56 years (range: 53-63), 4 patients were female, and 6 had brown/black skin **(Table 1)**. The average duration of disease was 13 years. Seven patients were on active treatment for sarcoidosis at the start of the study, including 5 taking prednisone **(Figure 1A)**. The average CSAMI activity score, assessed while taking the preceding therapeutic regimen, was 37 (range: 19-56).

**Figure 1.**
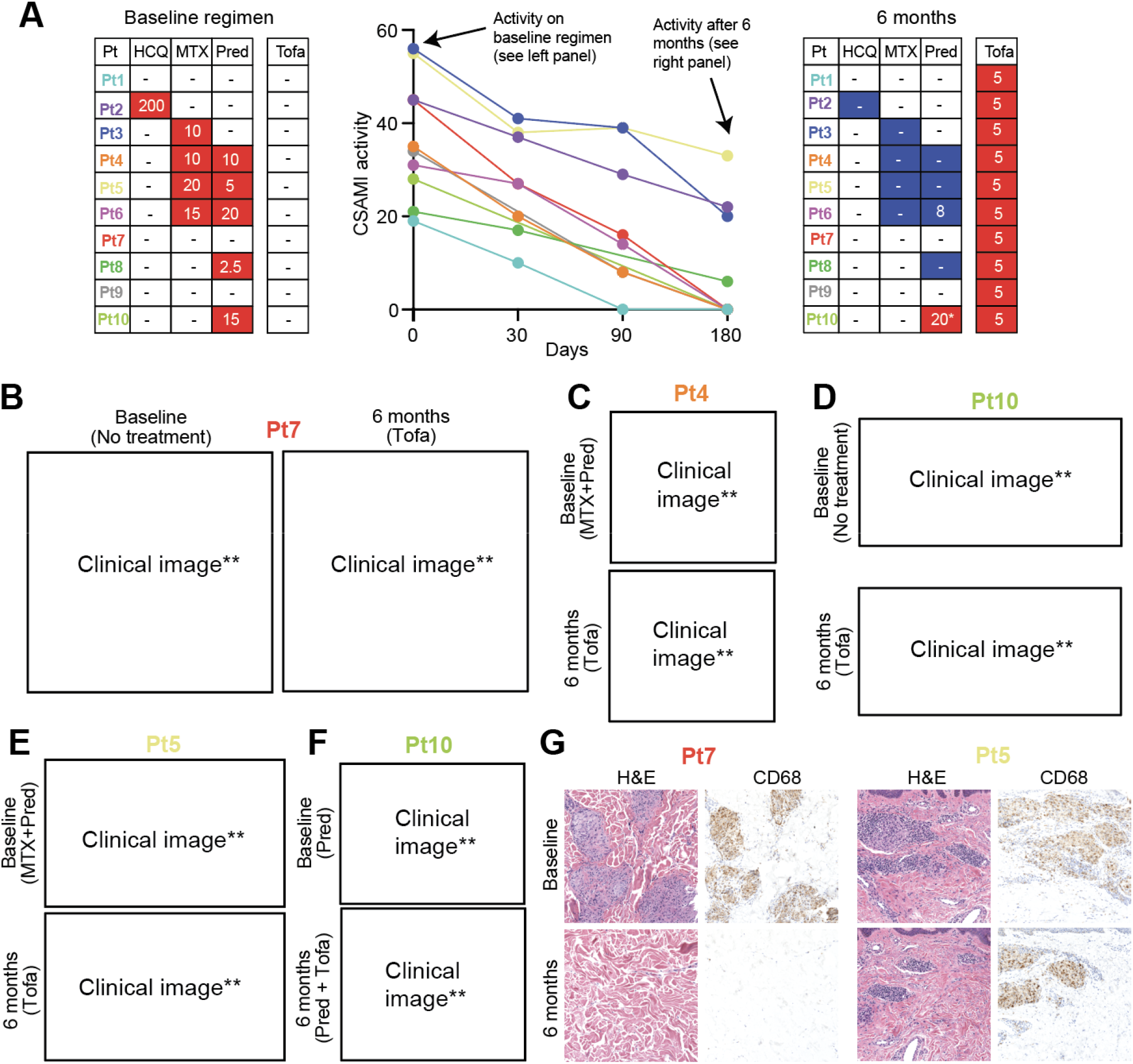
Tofacitinib treatment leads to improvement in cutaneous sarcoidosis. **(A)** Left panel: baseline treatment regimens for each patient, HCQ: hydroxychloroquine (dose shown as mg twice daily), MTX: methotrexate (mg weekly), Pred: prednisone (mg daily), and Tofa: tofacitinib (mg twice daily). Blue: discontinued/reduced dose during study. Middle panel: CSAMI activity scores over the study period. Right panel: treatment regimens after 6 months for trial patients, *prednisone increased due to worsening pre-existing Achilles tendonitis, not worsening of sarcoidosis. **(B)** Dermal papules/plaques and subcutaneous nodules of sarcoidosis before and after treatment. **(C)** Lupus pernio presentation of sarcoidosis before and after treatment. **(D)** Papules/plaques on the forearm of a patient before and after treatment. **(E)** Sarcoidal dactylitis before and after 6 months of treatment. **(F)** Lupus pernio presentation of sarcoidosis before and after treatment; significant scarring persisted in this patient. **(G)** Hematoxylin and eosin (H&E) stained and CD68 immunohistochemistry on skin biopsies from a representative complete responder (left panels) and partial responder (right panels). **Some potentially identifying patient information has been removed.

All patients had improvement in the CSAMI activity score after 6 months of tofacitinib, with an average reduction of 82.7% (range: 40%-100%) **(Figures 1A-F)**. Six patients had a compete response (defined as CSAMI activity score of 0). Post-inflammatory hyperpigmentation and/or scarring often persisted (e.g. **Figure 1F**).

A skin biopsy was performed before and after 6 months of treatment in 6 patients; of which 3 showed a complete response and 3 showed partial improvement. In patients with a complete clinical response, sarcoidal granulomas were no longer evident in skin. In those with a partial clinical response, biopsy of active areas showed persistence of granulomatous inflammation (**Figure 1G)**.

### Tofacitinib leads to improvement in pulmonary and cardiac sarcoidosis

Of the 10 patients, 8 had pulmonary involvement, and 1 had myocardial involvement **(Table 1)**. Whole body PET-CT was performed in these 9 patients and total lesion glycolysis (TLG) was determined to assess sarcoidosis activity. Baseline studies were performed while taking a stable preceding immunosuppressive regimen that included prednisone plus methotrexate in 3 patients, and prednisone or methotrexate monotherapy in 2 and 1 patients, respectively **(Figure 1A)**. A subsequent PET-CT after 6 months of tofacitinib was obtained. One follow-up study could not be interpreted due to nonadherence to the dietary preparation.

A decrease in TLG of ≥50% was noted in 5 patients, with complete or near complete resolution in 3 (>98% reduction in TLG) **(Figure 2A)**. One patient had essentially stable TLG, but did appear to have improvement in mild lymph node avidity below the detection threshold **(Figure 2B)**. Two patients had an increase in TLG, however; both were able to discontinue methotrexate and prednisone during the study and the change was not clearly clinically significant. Further, both experienced improvement in their skin (e.g. **Figure 2C)**. Examples of complete or near complete resolution in pulmonary **(Figure 2D-E)** and myocardial disease activity **(Figure 2F)** are illustrated.

**Figure 2.**
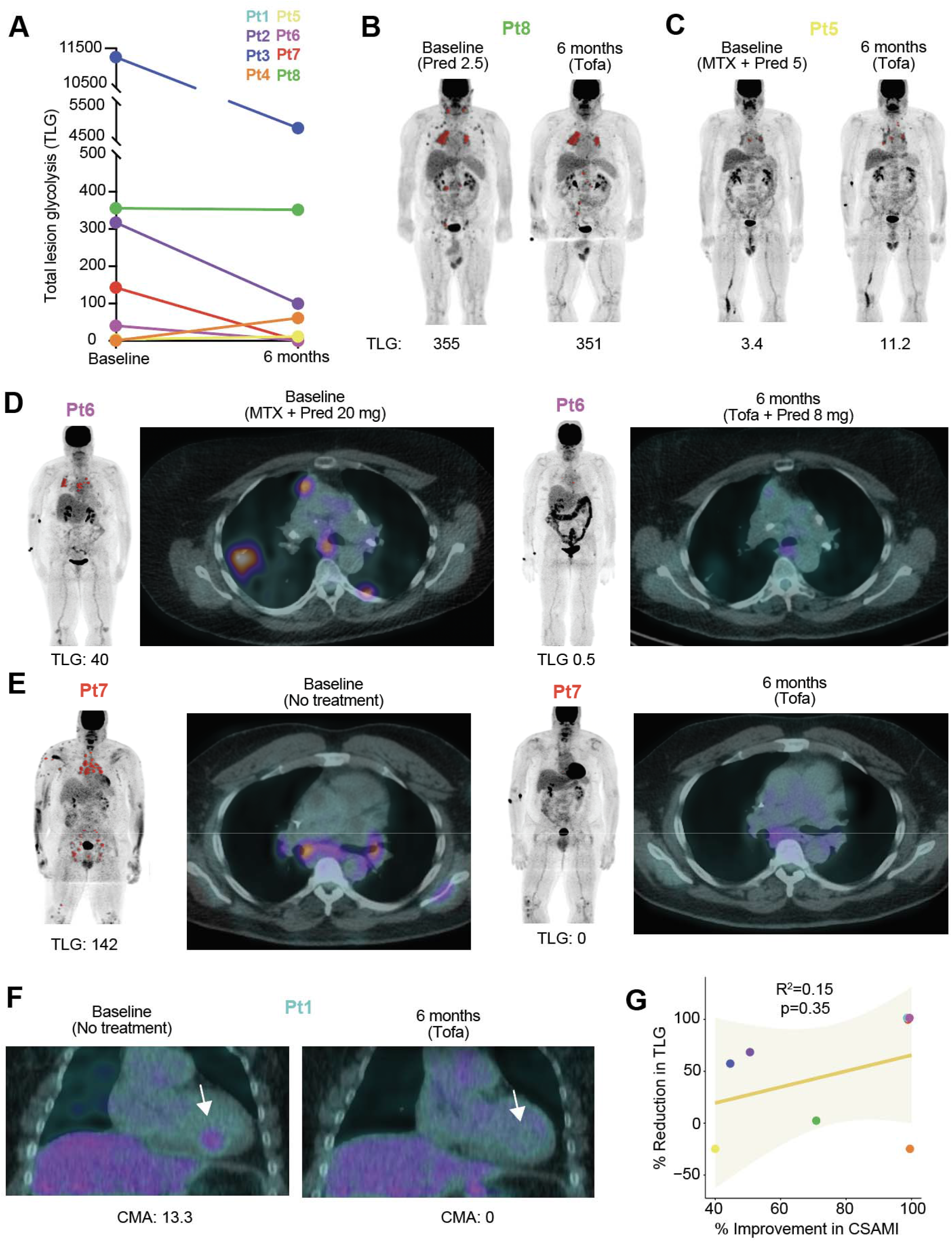
Tofacitinib treatment leads to improvement in pulmonary and cardiac sarcoidosis. **(A)** Change in total lesion glycolysis (TLG) during the study period. Baseline and 6-month treatment regimens as depicted in **Figure 1A. (B)** PET before and after 6 months of treatment in a patient with stable disease. **(C)** PET before and after 6 months of treatment in representative patient with increase in TLG. **(D-E)** PET and PET-CT (axial) before and after 6 months of treatment in patients with complete or near complete response. **(F)** PET-CT (coronal) before and after 6 months of treatment in a patient myocardial involvement of the inferior intraventricular septum (arrow), CMA: cardiac metabolic activity. **(G)** Scatterplot comparing change in cutaneous sarcoidosis and extra-cutaneous sarcoidosis. Depicted as regression line with 95% confidence interval (shared area) Cutaneous involvement shown as percent reduction in CSAMI during the study period. Extracutaneous involvement shown as percent reduction in TLG during the study period; for patients with increase in TLG during the study, worsening of 50% was arbitrarily assigned.

Of 5 patients taking prednisone at baseline, three were able to discontinue it entirely and one was able substantially decrease the dose. One patient with pre-existing, Achilles tendinopathy responsive to prednisone increased his prednisone dose during the study for this. All patients taking methotrexate were able to discontinue it. Several patients reported subjective improvement in confirmed or suspected nasal sinus involvement. A patient with a hoarse voice due to laryngeal involvement noted normalization of her voice.

In general, the skin tended to improve to a greater degree than internal organ involvement. There was weak, non-significant correlation between the degree of improvement in cutaneous and internal organ involvement **(Figure 2G)**. Definitive interpretation of this trend was limited by varying baseline treatment regimens.

In all 10 patients, the treatment regimen including tofacitinib led to disease improvement not achieved with the prior regimen. All 10 patients reported improved skin-related quality of life **(Figure S1)**. Tofacitinib was well tolerated, there were no significant or dose-limiting adverse events **(Table S1)**.

### IFN-γ produced by Th1 polarized CD4+ T cells activates lesional macrophages in sarcoidosis

In order to better understand the potential molecular targets of tofacitinib in sarcoidosis, single cell RNA sequencing (scRNA-seq) was performed on lesional skin from 3 of the patients prior to treatment and compared with normal skin from 3 healthy controls **(Table S2)**. 24034 cells were analyzed with a median of 4029 unique molecular identifiers. The data were visualized using uniform manifold approximation and projection (UMAP), revealing 37 clusters, corresponding to 11 major cell types **(Figure S2)**. Although transcriptional variation was noted in several cell types, when comparing sarcoidosis to normal, we focused our analysis on T cells and myeloid cells given the important role that these cell types play in granuloma formation^24^.

A total of 11236 T cells were clustered independently of the other cell types, revealing 14 unique clusters composed of helper CD4+ T cells (T_h_) (*CD4*+*FOXP3*-), regulatory T cells (*CD4*+*FOXP3*+), and CD8+ T cells (*CD8A*+) **(Figures 3A-B, S3)**. T_h_ cells were the most abundant T cell type in sarcoidosis consistent with prior observations, accounting for 70.2 % of T cells and were the focus of the remainder of the analysis.

**Figure 3.**
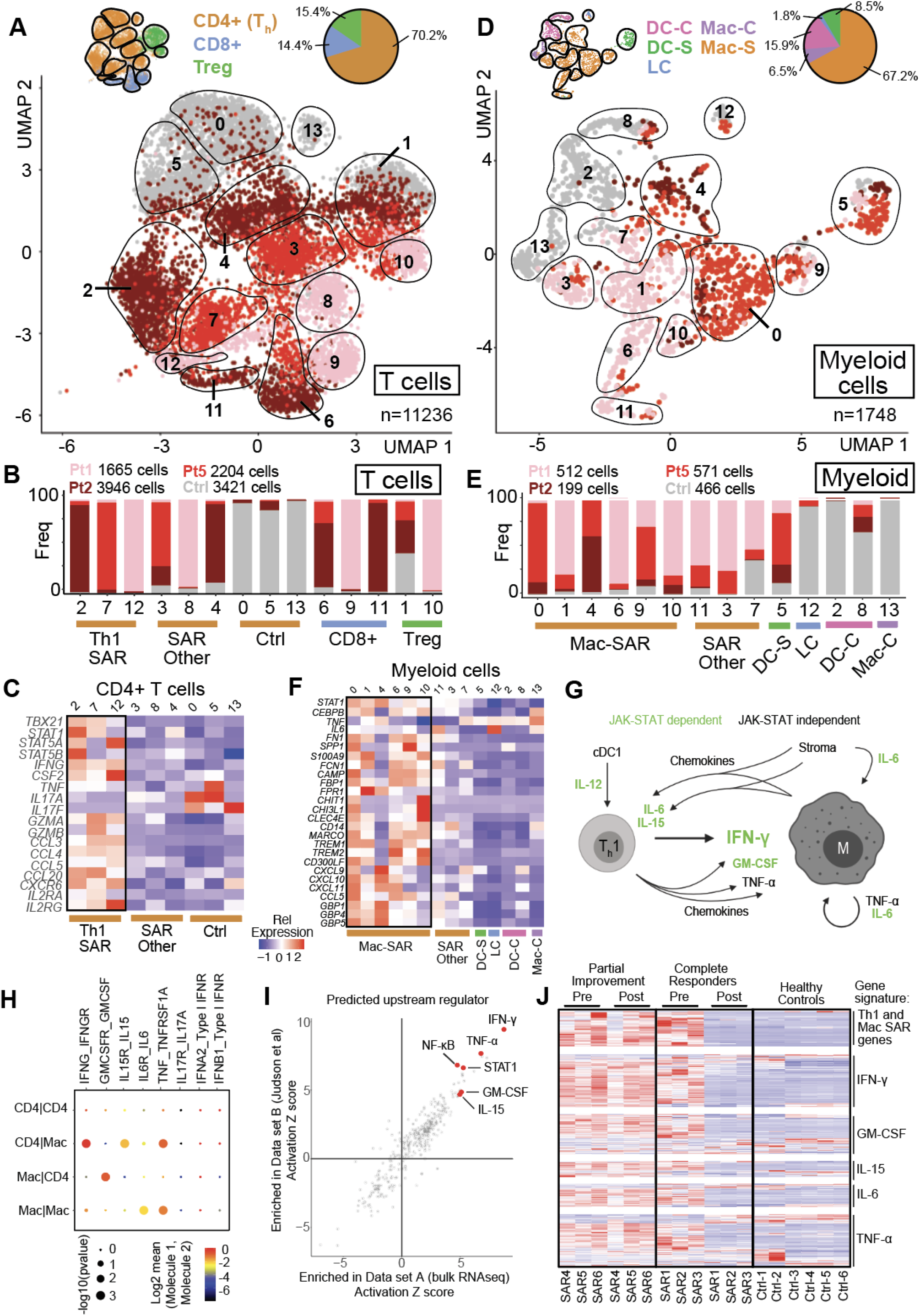
IFN-γ produced by Th1 polarized CD4+ T cells activates lesional macrophages in sarcoidosis. **(A)** UMAP projection of scRNA-seq data showing T cell clusters in sarcoidosis compared to healthy controls. **(B)** Histograms showing contribution of each condition (sarcoidosis: shades of red, grey: controls) to each T cell cluster. **(C)** Heatmap showing expression of selected transcripts in T cell clusters. **(D)** UMAP projection of scRNA-seq data showing myeloid cell clusters in sarcoidosis compared to healthy controls. **(E)** Histograms showing contribution of each condition (sarcoidosis: shades of red, grey: controls) to each myeloid cluster. **(F)** Heatmap showing expression of selected transcripts in myeloid clusters. **(G)** Summary of central cytokine and chemokine signals in sarcoidosis revealed by scRNA-seq. **(H)** Dot plot of cellphone DB receptor-ligand interaction analysis for select molecule and receptor pairs.**(I)** IPA analysis showing predicted upstream regulators in cutaneous sarcoidosis dataset A (RNAseq) and cutaneous sarcoidosis dataset B (Judson et al)^28^. **(J)** Heatmap showing expression of selected transcripts.

Differential gene expression and pathway (IPA) analysis were used to compare predominant T_h_ populations in sarcoidosis (clusters 2, 7, 12) to controls (clusters 0, 5). revealing a unique activated T_h_1 cluster in each sarcoidosis patient (T_h_1-SAR) that was not present in controls **(Figures 3B, S3)**. *IFNG* (IFN-γ) was by far the most differentially upregulated cytokine in T_h_1-SAR, which expressed high levels of canonical T_h_1 transcription factors (*TBX21, STAT1*). Increased expression of *CSF2* (GM-CSF) was also present. *TNF* (TNF-α) was expressed at modest levels and there was minimal IL-17 expression **(Figure 3C, S3)**. Chemokines CCL3, CCL4, and CCL5 were also highly upregulated **(Figure S3)**. Predicted upstream regulators of T_h_1-SAR included IL-6, IL-12, and IL-15 **(Figure S3)**.

We next analyzed myeloid clusters. A total of 1748 myeloid cells were analyzed, revealing 14 clusters which included both macrophages and dendritic cells (DCs) **(Figure 3D-E, S4)**. Macrophages were abundant and somewhat diverse in sarcoidosis (**Figure 3E**). Expression patterns among predominant macrophage populations in sarcoidosis (Mac-SAR, clusters 0, 1, 4, 6, 9, 10) were compared to control clusters (clusters 2, 8, 13). Mac-SAR clusters expressed high levels of transcription factors *STAT1* and *CEBPB*, activation/effector molecules (*CHIT1, CHI3L1, TREM1, TREM2*), T cell chemokines (*CXCL9, CXCL10, CXCL11*), and interferon signature genes (*GBP1, GBP4, GBP5*), among others **(Figure 3F)**. A population of activated DCs with a cDC1 phenotype that produced IL-12 were present almost exclusively in the sarcoidosis samples **(Figure S4)**. IFN-γ was the most highly significant inferred upstream cytokine regulator of Mac-SAR based on their transcriptional profile **(Figure S4)**.

Together, these data strongly implicated IFN-γ derived from an aberrant T_h_1 response as a key driver of sarcoidosis pathology, consistent with the conserved role of IFN-γ in classical macrophage activation and granuloma formation ^25,26^. Through these and other analyses (below) we identified other cytokine signals including GM-CSF, IL-15, IL-6, TNF-α, which we hypothesize play a secondary/reinforcing role **(Figure 3G)**. Proposed cytokine and chemokine interactions among T cells and macrophages were further supported by a bioinformatic analysis of receptor-ligand interactions (Cellphone DB)^27^ **(Figure 3H)**. Interestingly, IL-15 and IL-6 were produced predominantly by fibroblasts and endothelial cells **(Figure S5)**. Of these signals IFN-γ (JAK1/2), GM-CSF (JAK2), IL-6 (JAK1/2), and IL-15 (JAK1/3) are expected to be inhibited by tofacitinib.

### Improvement correlates with suppression of IFN-γ and other cytokine activity in tissue

In 6 patients for whom matched skin biopsy specimens were available **(Table S3)**, bulk RNA sequencing was performed on lesional skin before and after tofacitinib. This data was compared to skin from 6 healthy controls and further contextualized using an independent gene expression data set consisting of 15 cutaneous sarcoidosis biopsies and 5 controls (Judson et al)^28^. IPA was performed to compare sarcoidosis to controls in our data and the Judson data and showed that an IFN-γ transcriptional signature was the most significant change in both data sets. IL-15, IL-6, GM-CSF, and TNF-α were also among the most significant predicted regulators **(Figure 3I)**.

Analysis of pre- and during-treatment skin biopsies from trial patients showed that in those achieving complete response, there was near-complete suppression of T_h_1-SAR and Mac-SAR genes (from scRNA-seq experiments) and an IFN-γ response signature **(Figure 3J)**. Whereas, in patients with partial improvement, T_h_1-SAR, Mac-SAR genes and the IFN-γ response signature were still detected. GM-CSF, IL-15, IL-6, and TNF-α response signatures also correlated somewhat with response patterns, albeit not as closely as IFN-γ, as assessed using principal component analysis **(Figures 3J, S6)**. Overall, the bulk expression data was highly consistent with the scRNAseq data and further implicated IFN-γ as a key driver of sarcoidosis.

### Tofacitinib reduces cytokine, chemokine, and macrophage activation marker levels in plasma

We used high throughput proteomics to study plasma from 9 patients in whom matched samples were available before and after 6 months of tofacitinib and compared these with 11 healthy controls **(Table S4)**. Of the 1536 proteins analyzed with this approach, we found IFN-γ, IL-6, and TNF-α to be among the most upregulated in sarcoidosis at baseline relative to controls **(Figure 4A)**. IL-15 was more modestly increased (GM-CSF was not part of this panel). Further, we found a significant correlation between levels of proteins in plasma and mRNA expression in cutaneous sarcoidosis, suggesting upregulation of several of these markers in plasma was a direct reflection of disease activity **(Figure 4B)**.

**Figure 4.**
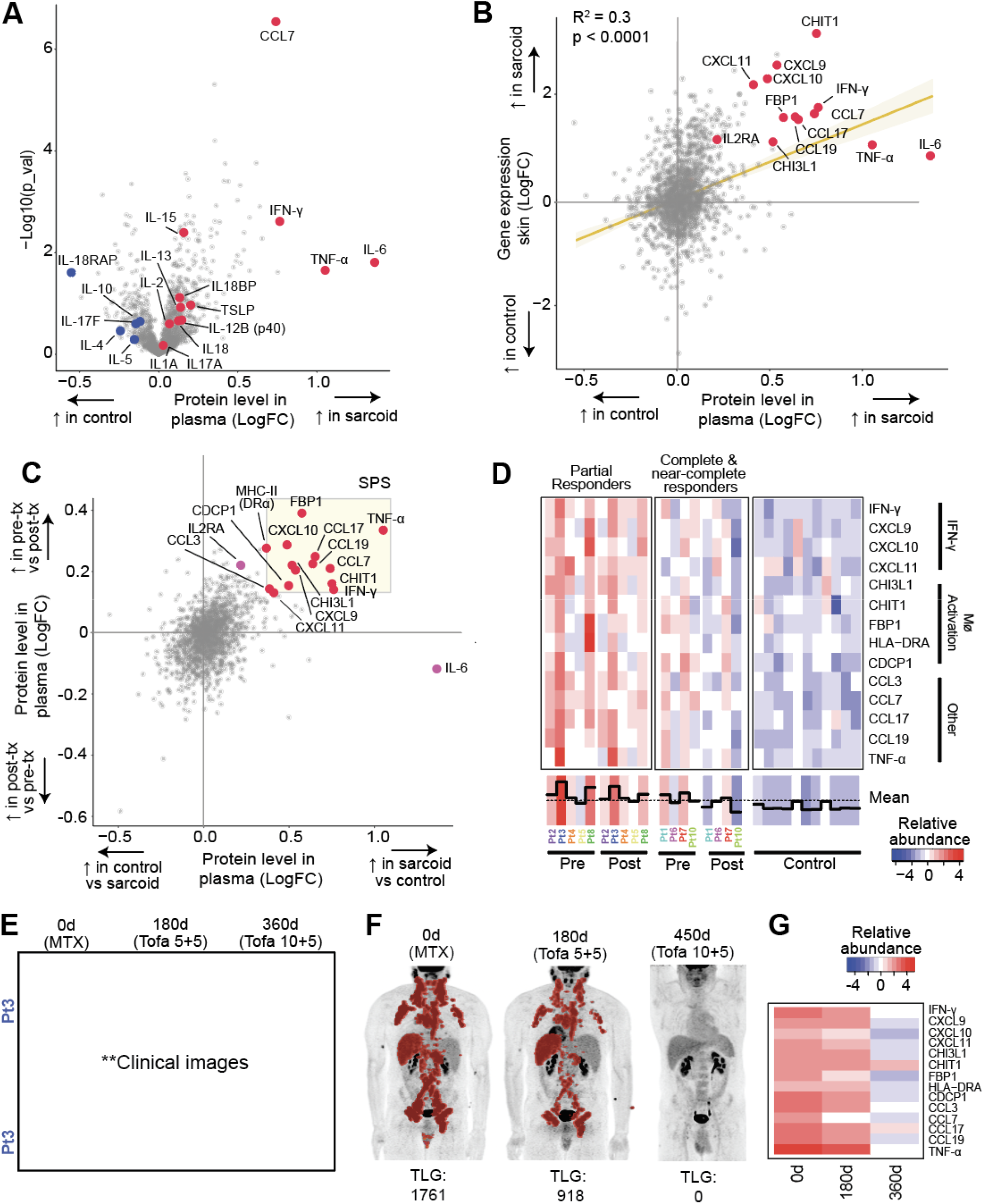
Tofacitinib reduces cytokine and chemokine levels and markers of macrophage activation in plasma. **(A)** Relative abundance of proteins in sarcoidosis plasma compared to controls. **(B)** Relative abundance of plasma proteins in sarcoidosis compared to controls (x-axis) compared to mRNA levels in skin of sarcoidosis compared to controls (y-axis). Regression line with 95% confidence interval (shaded area) shown. **(C)** Relative abundance of proteins in sarcoidosis plasma compared to controls (x-axis) plotted against relative abundance of proteins in sarcoidosis plasma at baseline (pre-tx) compared to after 6 months of tofacitinib (post-tx) (y-xais). The most consistently and highly differentially abundant proteins were used to create the sarcoidosis plasma signature (SPS) shown in the shaded box. **(D)** Heatmap showing levels of SPS proteins in trial participants (before and after tofacitinib) and healthy controls, grouped by response pattern **(E)** Photographs of scalp and scrotal sarcoidosis. Shown at baseline, after 6 months (180d) of tofacitinib at 5 mg twice daily, and after 12 months (360d) of tofactinib 10 mg in the morning and 5 mg at night. **(F)** PET scans of the patient and E before tofacitinib, after 6 months (180d) at 5 mg twice daily, and after 14 months (420d) at 10 mg in the morning and 5 mg at night. **(G)** SPS levels corresponding to timepoints in **(E)**. **Some potentially identifying information has been removed.

Next, we overlaid 1) proteins upregulated in sarcoidosis plasma at baseline versus controls with 2) proteins modulated by tofacitinib therapy in sarcoidosis plasma, and in doing so found a 14-protein signature or sarcoidosis that we termed, sarcoidosis plasma signature (SPS). The SPS included IFN-γ, IFN-γ targets (CXCL9, CXCL10, CXCL11), macrophage activation proteins (CHIT1, CHI3L1, and FBP1) and TNF-α **(Figure 4C)**. We then evaluated whether differences in SPS profiles correlated with response to tofacitinib. To do so, we compared SPS levels in the 4 best responders (complete/near complete response) to the other 5 patients (partial improvement) **(Figure 4D)**. We found that the best responders tended to have lower baseline SPS activity and tofacitinib resulted in relative normalization of SPS to levels comparable to healthy controls. In contrast, the other patients tended to have higher baseline SPS activity (or were heavily immunosuppressed when the baseline samples were collected, e.g. Pts 4,5) with SPS levels tending to remain elevated above those seen in healthy controls even with tofacitinib **(Figure 4D)**.

### Evidence for dose-dependency of the response to tofacitinib in sarcoidosis

Given that relatively higher baseline SPS levels in plasma and persistent IFN-γ activity in skin were associated with partial improvement on tofacitinib, we hypothesized that in some patients, a higher dose of tofacitinib might be required. In order to explore this further, a single patient with partial improvement in cutaneous, pulmonary, and lymph node sarcoidosis after 6 months of tofacitinib 5 mg twice daily during the trial **(Figure 4E-F)** was subsequently treated with a higher dose of tofacitinib (10 mg in the morning and 5 mg at night) obtained through insurance after the trial. After an additional 6 months of treatment at the higher dose, there was complete clearance of his skin and after 8 months there was resolution of pulmonary and diffuse lymph node FDG-avidity **(Figure 4E-F)**. A relative normalization of the SPS signature was also observed on the higher dose **(Figure 4G)**. These data suggest that a higher dose of tofacitinib might be required in patients with higher baseline disease activity.

## Discussion

In this open-label trial of tofacitinib, we demonstrate efficacy in 10 sarcoidosis patients with cutaneous involvement. In all 10 patients, disease control with a tofacitinib-based regimen was superior to the patients preceding immunotherapeutic regimen, particularly for skin involvement. Four of five patients entering the study taking prednisone were able to discontinue or significantly reduce the dose.

Our mechanistic evaluation suggested that IFN-γ is a key driver of sarcoidosis and is a critical cytokine targeted by tofacitinib with effective treatment. This observation is consistent with prior work showing that IFN-γ is elevated in circulation and in tissues of sarcoidosis patients and correlates with disease activity^29-32^, and makes teleological sense given the fundamental role of IFN-γ in classical macrophage activation, granuloma formation and protection against *Mycobacterium tuberculosis*^25,26^. Tofacitinib appears to provide an effective means of suppressing IFN-γ, which signals via JAK1/2, in sarcoidosis.

We also found that the activity of other cytokines including GM-CSF (JAK2), IL-15 (JAK1/3), IL-6 (JAK1/2), and TNF-α (JAK-independent) are also evident in sarcoidosis. GM-CSF has been shown to promote the differentiation of monocytes into inflammatory macrophages in autoimmunity^33^, IL-15 can re-enforce CD4+ T cell effector responses^34^, and IL-6 is an additional proinflammatory cytokine implicated as a potential treatment in sarcoidosis^35^. Although we do not exclude a role for TNF-α in sarcoidosis, we do implicate these additional cytokines, which may also need to be inhibited for maximal response to therapy. A potential advantage of JAK inhibition (compared to TNF-α inhibition) is the simultaneous, direct inhibition of multiple cytokines. Furthermore, we observe a reduction in TNF-α production and activity with tofacitinib, suggesting that TNF-α production may occur, at least in part, downstream of these JAK-STAT dependent cytokines.

While all patients improved, some patients still had disease activity on tofacitinib. This was associated with incomplete suppression of IFN-γ activity, suggesting that higher doses might be required in some patients. Indeed, in a single patient, a higher dose of tofacitinib led to remission of cutaneous and internal organ sarcoidosis. This is consistent with a prior case report, showing a better response of widespread sarcoidosis to tofacitinib 10 mg twice daily than to 5mg twice daily^16^. Moving forward, improved suppression of IFN-γ activity could be achieved through either a higher dose of tofacitinib in some patients, or, potentially evaluation of more targeted JAK inhibitors, such as a JAK1- or JAK1/2-specific inhibitor.

Together, these data are promising and support further evaluation of JAK inhibition in the treatment of sarcoidosis.

## Data Availability

scRNA-seq, bulk RNA-seq, and proteomic data have been uploaded to Gene Expression Omnibus (Accession number pending).

## Acknowledgements

The authors would like to thank G. Wang and the Yale Center for Genome Analysis as well as M. Zhong and the Yale Stem Cell Center sequencing core for assistance with scRNA-seq and bulk RNA-seq experiments, respectively. WD is supported by a Career Development Award from the Dermatology Foundation. CR is supported by research funding from the Chest Foundation and NIH/NHLBI (1K08HL151970-01). scRNA-seq, bulk RNA-seq, and proteomic data have been uploaded to Gene Expression Omnibus (Accession number pending).

## SUPPLEMENTARY MATERIAL

### SUPPLEMENTARY METHODS

#### Patient selection and informed consent

Potentially eligible patients with sarcoidosis were identified in Yale Medicine clinics (dermatology, pulmonology, and cardiology). Inclusion criteria included: 1) a diagnosis of cutaneous sarcoidosis with supportive skin biopsy, 2) Cutaneous Sarcoidosis Activity and Morphology Instrument (CSAMI)(1) activity score of 10 or greater, and 3) taking a stable dose of oral immunomodulatory regimen for at least 3 months, with no plans to change the regimen over the subsequent 6 months. Patients less than 18 years of age, a history of chronic, untreated infections, and malignancy were excluded. Most patients also had internal organ involvement, but it was not used as a criterion for eligibility. Full inclusion/exclusion criteria are listed in the study protocol. Patients provided written informed consent prior to any study related interventions. The study was registered with clincaltrials.gov (NCT03910543).

#### Clinical dermatologic evaluation and specimen acquisition

The clinical extent of cutaneous sarcoidosis was quantified using the validated Cutaneous Sarcoidosis Activity and Morphology Instrument (CSAMI)(1), which quantifies disease activity (representing active inflammation) and damage (representing tissue damage/destruction) separately. Only the activity portion of the CSAMI instrument was utilized for this study. Two 4mm punch biopsies of lesional skin were obtained from each patient, prior to initiation of tofacitinib (while taking the preceding immunomodulatory treatment regimen) and again after 6 months of tofacitinib. In patients where disease activity remained on tofacitinib, a lesion that still appeared active was biopsied. In patients with an apparent complete clinical response to tofacitinib, a representative, previously active area of skin was biopsied. In patients with primarily facial involvement, the research biopsies were not mandatory. A portion of the biopsy tissue from each patient was placed in RNAlater reagent (Qiagen) and snap frozen for downstream analyses (see below) and another portion was fixed in 10% buffered formalin and embedded with paraffin for histologic analysis (see below). In 3 patients, a third biopsy was obtained from lesional skin prior to initiation of tofacitinib and was used for single cell RNA sequencing (see below). Skindex-16 is a validated, skin-related quality of life tool (2). Skindex-16 was administered at baseline and after 6 months of tofacitinib therapy.

#### FDG-PET/CT with Rubidium-82 Myocardial Perfusion Imaging Acquisition, Processing, and Analysis

FDG-PET imaging was performed in the morning following a one-day high fat/low carbohydrate diet followed by an overnight fast of greater than 12 hours as previously described(3). Dietary preparation instructions were provided in writing to the patient, along with telephone contact by clinical staff 48-72 hours prior to the study.

Both the cardiac Rubidium-82 (Rb-82) and whole-body FDG-PET imaging were performed on a GE Discovery D690 PET/CT scanner. Low-dose CT images (120 kV, 50 mA for BMI <39, modulated at 50-150 mA for BMI >39) for the purposes of attenuation correction were performed before Rb82 and FDG imaging sequences.

Resting ECG-gated dynamic Rb-82 PET imaging were performed using 20-35 millicurie (mCi) of Rb82 consistent with established clinical guidelines and our previous publications(4). Following Rb-82 imaging, the patient was injected with FDG (8-10mCi), followed by a 60-minute waiting period during which the patient relaxed or read in a quiet room. Whole-body FDG images were acquired for 2.5 minutes per bed position from cranial apex to knee, and 2 minutes per bed position from knees to toes. A separate, 8-minute single bed position cardiac acquisition was then performed.

Resting Rb-82 perfusion imaging was processed and interpreted visually with 4DM (Invia). Quantitative and visual analysis of cardiac FDG uptake was performed on the AW software platform (GE) on the dedicated 3D cardiac acquisition window (single 47 slice bed position) and reported as previously described(5, 6). (Quantitative and visual analysis of extra-cardiac FDG update is described below.) When present, the relationship between the location of inflammation and perfusion defects was characterized. The maximum standardized uptake value (SUV) in the heart was used to represent the peak level of inflammation. The volume of inflamed myocardium, cardiac metabolic volume (CMV), was identified as myocardium exceeding a SUV threshold that was derived for each patient by multiplying the left ventricular (LV) blood pool (background) activity by 1.5 as previously described(3). Cardiac metabolic activity (CMA) was defined as the CMV multiplied by the average SUV of that volume.

At the time of six-month follow up imaging, patients were provided with their written diet log from the initial scan, and instructed to replicate the same high-fat/low carbohydrate diet and fast duration as closely as was feasible. Follow up scans were performed using the same protocol and PET scanner.

The sarcoidosis metabolic lesion burden at baseline and post-treatment follow-up was determined using the whole-body PET segmentation tool LesionID® (MIM Software Inc, Cleveland, Ohio, U.S.A.). This tool allows semi-automatic determination of maximal and mean standardized uptake values (SUV) (SUV_max_ and SUV_mean_, respectively), mean tumor volume (MTV, cm^3^), and total lesion glycolysis (TLG, product of SUV_mean_ and MTV) of individual sarcoidosis lesions, and subsequent automatic calculation of MTV and TLG for the whole body. In the absence of a dedicated inflammatory lesion lexicon, the oncology lexicon was used to define metabolic parameters for sarcoidosis burden assessment.

The initial automatic segmentation (lesion contouring) performed by the software was based on a predefined threshold which was defined by placing a 3 cm diameter sphere in the right lobe of the liver. The threshold value was determined using 1.5 * liver SUV_mean_ + 2 * liver standard deviation, as defined by PET Response Criteria in Solid Tumors (PERCIST)(7). All voxels above this value were segmented and volumetrically separate regions were made into single volumes of interest (VOIs). The automatic segmentation was subsequently corrected by consensus of 3 readers, a dual-boarded radiologist and nuclear medicine physician (15 years of experience) (D.P.), a board-certified cardiologist (B.D.Y.), and a board-certified dermatologist (W.D.). During the correction process, the readers rejected false-positive lesions (mostly attributed to physiological uptake or pathologic uptake deemed sarcoidosis-unrelated) and made any necessary additional edits (modifying segmentation to avoid physiological uptake). All regions were combined into a total lesion burden VOI that encompassed all approved lesions for that timepoint.

All lesions were tracked across time at the follow-up study. On follow-up scan, we placed a liver reference region in the right lobe of the liver in order to calculate the threshold value specific to this timepoint. Tracked lesions were transferred from the baseline PET/CT to the follow-up PET/CT via a rigid fusion and redefined automatically using PET Edge+®, a hybrid intensity and gradient-based tool(8). We adjusted the tracked lesion VOIs as described above, if necessary. Next, similarly to the baseline timepoint, the rest of the image volume, not including the tracked lesions, was segmented with the PERCIST value specific to the follow-up timepoint. We followed the same steps as described above on the baseline timepoint to approve and finalize a total lesion burden VOI for the follow-up timepoint.

The software then calculated all metabolic parameters from the total lesion burden VOI based on final lesion contours created by semi-automatic segmentation and individually tracked lesions were compared across both timepoints. Baseline and follow-up studies were processed simultaneously for immediate automatic determination of treatment-related changes in the individual patients.

#### Management of other immunosuppression during the study period

Most patients were on therapy for their sarcoidosis at the outset of the study. Patients had the option to continue, change the dose, or discontinue other immunosuppressive medications at the outset, or during the trial. One patient was taking hydroxychloroquine and it was discontinued upon initiation of tofacitinib. Four patients were taking methotrexate at the outset, all elected to discontinue this medication upon initiation of the tofacitinib. Five patients were taking prednisone. Throughout the 6-month study period, patients were permitted to taper and/or discontinue the prednisone based on their symptoms and physical examination findings and the investigators’ discretion. In one patient, prednisone was increased during the study period related to increased pain associated with pre-existing Achilles tendonopathy.

#### Statistical Analysis

Statistical analysis was performed using GraphPad Prism 9. Simple linear regression analysis and P value determination was performed. The 95% confidence interval bands were plotted in Prism. Line graphs and histograms were also plotted in Prism.

#### Histology and immunohistochemistry

Histology and immunohistochemistry (IHC) were performed on formalin-fixed, paraffin-embedded sections using standard methods. For IHC, antigen retrieval was performing using citrate buffer (pH 6.0) (Life Technologies). The CD68 antibody was obtained from Dako (PG-M1). Primary antibody binding was detected and visualized using ImmPRESS peroxidase reagent kit (Vector) and diaminobenzidine (DAB) substrate (Vector).

#### RNA extraction and sequencing

Skin biopsy tissue stored in RNAlater reagent (Qiagen) was thawed on ice. The tissue was homogenized using a rotor-homogenizer (PowerGen 125, Fisher Scientific). Total RNA was extracted using the RNeasy Fibrous Tissue Mini Kit (Qiagen) according to the manufacturer’s instructions. Two healthy controls were also included **(Table S2)**. RNA was submitted to the Yale Stem Cell Center and complementary DNA (cDNA) libraries were generated and 100-base pairs paired-end sequencing was performed on an Illumina HiSeq 4000 at Yale Stem Cell Center.

Bulk RNA-seq data was initially processed using Partek Flow software (Version 9.0, build 9.0.20.0510). Burroughs-Wheeler Aligner (BWA, Version 0.7.15) was used to align the reads against hg38 (hg38_refseq_16_02_02 was used for quantification). Additional, previously published healthy control skin RNAseq samples run on the same platform were also included (GEO Accession number GSE122592)(9). Reads were normalized by counts per million (CPM). Further processing to identify differentially expressed genes between control, pre-treatment, and during treatment groups was carried out using DESeq2. Target gene transcript lists were downloaded from Gene Set Enrichment Analysis Molecular Signatures Database or Ingenuity Pathway Analysis(10).

#### Single cell RNA sequencing (scRNA-seq)

4 mm punch biopsies of skin were obtained from 3 patients with sarcoidosis and 3 healthy volunteers **(Table S3)**. Biopsies were immediately biopsied and placed in to Dispase II (10 mg/mL) (Sigma) in RPMI with 2% FBS for 45 minutes at 37°C with shaking at 125 RPM. The dispase solution was placed on ice. The tissue was removed from the dispase solution, minced in a sterile plate with a scalpel, and then subsequently incubated in Liberase TH (0.5 mg/mL) (Sigma) in RPMI with 2% FBS for 45 minutes at 37°C with shaking at 125 RPM. The dispase solution was placed on ice. After incubation in the liberase and the dispase solution was combined (to recover any additional cells from the dispase) and the mixture was triturated and filtered through a 100 μM sterile filter. Cells were washed with RPMI + 2% FBS and RBCs were lysed with ACK Lysing Buffer (Lonza) and washed again in RPMI + 2% FBS. Cells were stained with LIVE/DEAD Red viability dye (ThermoFisher) in RPMI + 0.1% FBS. Viable single cells were isolated using a BD FACS Aria cell sorter and sorted into cold RPMI + 2% FBS. Up to 10,000 cells (depending on the yield) were loaded onto a 10X Chromium Controller. Single cell libraries were prepared according to the manufacturer’s instructions by the Yale Center for Genome Analysis (YCGA). Sequencing was performed with one sample/library per lane on an Illumina Hiseq 4000 by YCGA.

#### scRNA-seq data analysis

Cellranger software (v3.1.0) was used to process the data. Cellranger mkfastq was used for processing raw files into fastq files, and cellranger count was used to align reads to hg38, filter the reads and generate a cell-by-gene matrix for each library. Libraries were aggregated using cellranger aggr without normalization to generate a single cell-by-gene matrix. The Seurat R package (v3.2.0) was used for further analyses. Droplets with ≤ 100 expressed genes were removed from the matrix. The NormalizeData command with a scaling factor of 10,000 was used to normalize counts. ScaleData was used to regress the data against the number of transcripts and center gene expression values. Principle component analysis (PCA) was performed using RunPCA. Cells were clustered using FindNeighbors and FindClusters. Clusters consisting of cells with low/null expression of *GAPDH* (non-cells) were removed from further analysis using the SubsetData command, resulting in 24,034 cells for analysis. The data was visualized by performing Uniform Manifold Approximation and Projection (UMAP) dimensional reduction.

Cell-type assignments for each cluster were determined using canonical markers. For immune cell subsets, cell type assignments for each cluster were verified by comparing with ImmGen datasets(11). T cell and myeloid cell clusters were subsetted and re-analyzed separately using the approach described above. Stacked histogram plots were generated using ggplot2 (v3.3.3), ggrepl (v0.9.1) and dplyr (v1.0.3) were used to generate dot plots.

#### Pathway analysis, Cellphone DB, and data visualization

Ingenuity Pathway Analysis (IPA, Ingenuity Systems QIAGEN, Content version: 51963813) was used to perform core analysis on both bulk RNA-seq data and scRNA-seq data. Canonical pathways, upstream regulator and causal network analyses were utilized to compare differentially expressed gene lists generated using Seurat (scRNA-seq) or Partek Flow (bulk RNAseq). ggplot2 v3.2.1, ggrepl (v0.8.2) and dplyr (v1.0.2) were used to visualize the results by plotting Z-score versus p-value. Selected genes or pathways were labeled at the investigators’ discretion. Heatmaps were generated using gplots (v3.0.4) heatmap.2 function. Raw values were scaled using the scale function. Manhattan clustering was utilized. For Cellphone DB (version 2.1.4), normalized counts and meta data were exported from Seurat and imported into CellphoneDB. The statistical_analysis command was used. Dotplots were generated using the dot_plot command.

#### Multiplex soluble protein analysis of plasma

Blood samples were collected from participants at baseline and again after 6 months of therapy. Samples were compared to 11 healthy, roughly age matched controls **(Table S4)**. Blood was collected in EDTA-coated tubes and centrifuged; the buffy coat was removed and stored separately from the plasma, which was aliquoted and stored at - 80°C. Plasma was analyzed using the Olink Explore 1536 panel. This is a multiplexed panel that quantifies 1536 protein analytes simultaneously using a proximity extension approach. Detection of individual proteins requires binding by two matched, oligo-barcoded antibody pairs for each target. When both antibodies are bound, the oligo tags come in close proximity to each other and hybridize forming a unique template for DNA polymerase-dependent extension. After PCR amplification, next generation sequencing is performed and processed by Olink (Boston, Massachusetts). Log-fold changes and p values were calculated for pre- vs post-treatment and pre-treatment vs healthy. The data was then imported into R to generate heatmaps and volcano plots using ggplot2 (v3.3.3), ggrepl (v0.9.1), gplots (v3.1.1) and dplyr (v1.0.3).

## SUPPLEMENTARY TABLES

**Table S1.** Summary of adverse events. *This was a chronic, pre-existing condition. The Achilles was not significantly PET avid arguing against it representative a specific manifestation of sarcoidosis. **Patient requested this approach as it had worked well for him in the past.

| Adverse event | Incidence | Outcome | Serious AE? |
| --- | --- | --- | --- |
| SARS-CoV-2 infection | 2 | resolved, inpatient care not required | no |
| Acute sinusitis | 1 | treated with oral antibiotics | no |
| Lymphopenia | 1 | transient | no |
| Elevated LFTs | 1 | transient | no |
| Hyperlipidemia | 1 | statin initiated | no |
| Weight gain | 1 | persisted | no |
| Worsening Achilles tendonopathy pain* | 1 | Treated with prednisone** | no |

**Table S2.** Samples used for scRNA-seq experiments.

| **Healthy controls** |  |  |  |
| --- | --- | --- | --- |
| Sample | Approx age | Sex | Location |
| 1 | 66-70 | F | arm |
| 2 | 36-40 | F | arm |
| 3 | 36-40 | M | arm |
| **Sarcoidosis patients** |  |  |  |
| Sample | Age | Sex | Location |
| Pt1 | 61-65 | F | arm |
| Pt2 | 51-55 | F | arm |
| Pt5 | 56-60 | M | arm |

**Table S3.**
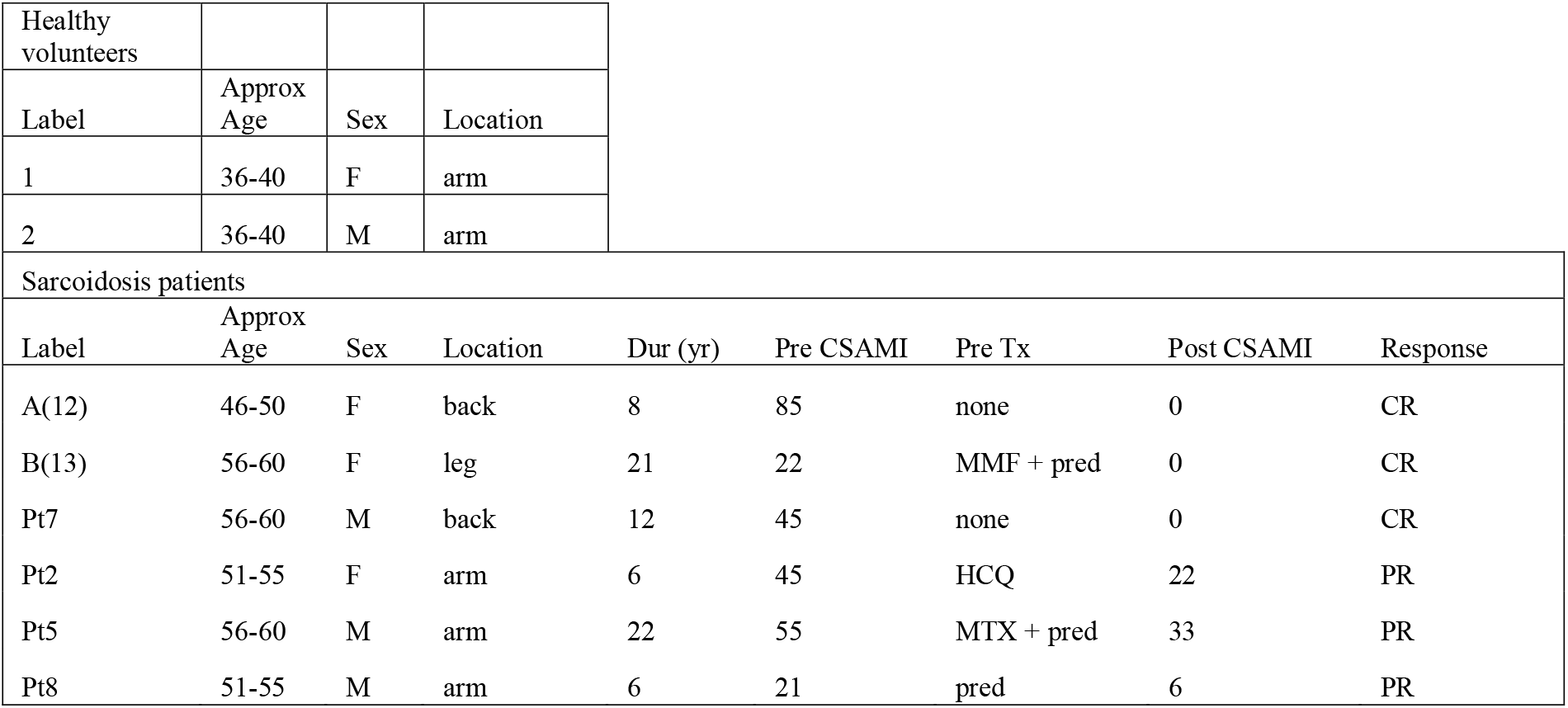
Samples used for bulk RNA-seq experiments. HCQ: hydroxychloroquine, pred: prednisone, MTX: methotrexate, MMF: mycophenolate. CSAMI: cutaneous sarcoidosis activity and morphology instrument activity score. Pre-Tx: baseline treatment regimen on which the biopsy was obtained. Post-Tx: obtained while taking tofacitinib 5 mg twice daily.

**Table S4.** Characteristics of control samples used for plasma proteome analysis (Olink).

| Healthy volunteers |  |  |
| --- | --- | --- |
| Label | Approx.<br>Age | Sex |
| Healthy 1 | 71-75 | F |
| Healthy 2 | 61-65 | M |
| Healthy 3 | 56-60 | F |
| Healthy 4 | 51-55 | F |
| Healthy 5 | 51-55 | F |
| Healthy 6 | 51-55 | F |
| Healthy 7 | 51-55 | F |
| Healthy 8 | 56-60 | M |
| Healthy 9 | 56-60 | F |
| Healthy 10 | 51-55 | F |
| Healthy 11 | 56-60 | F |

**Table S5.**
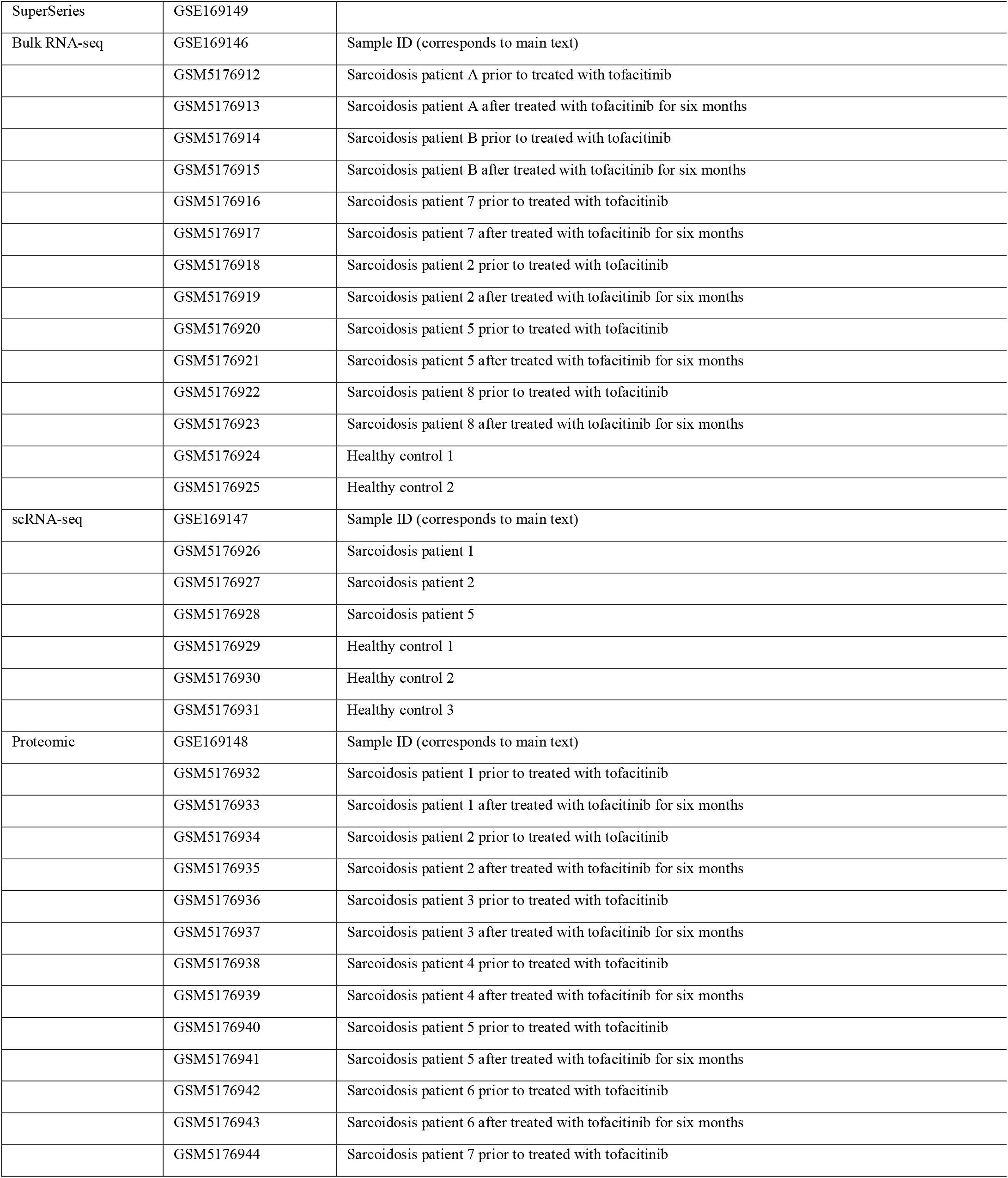
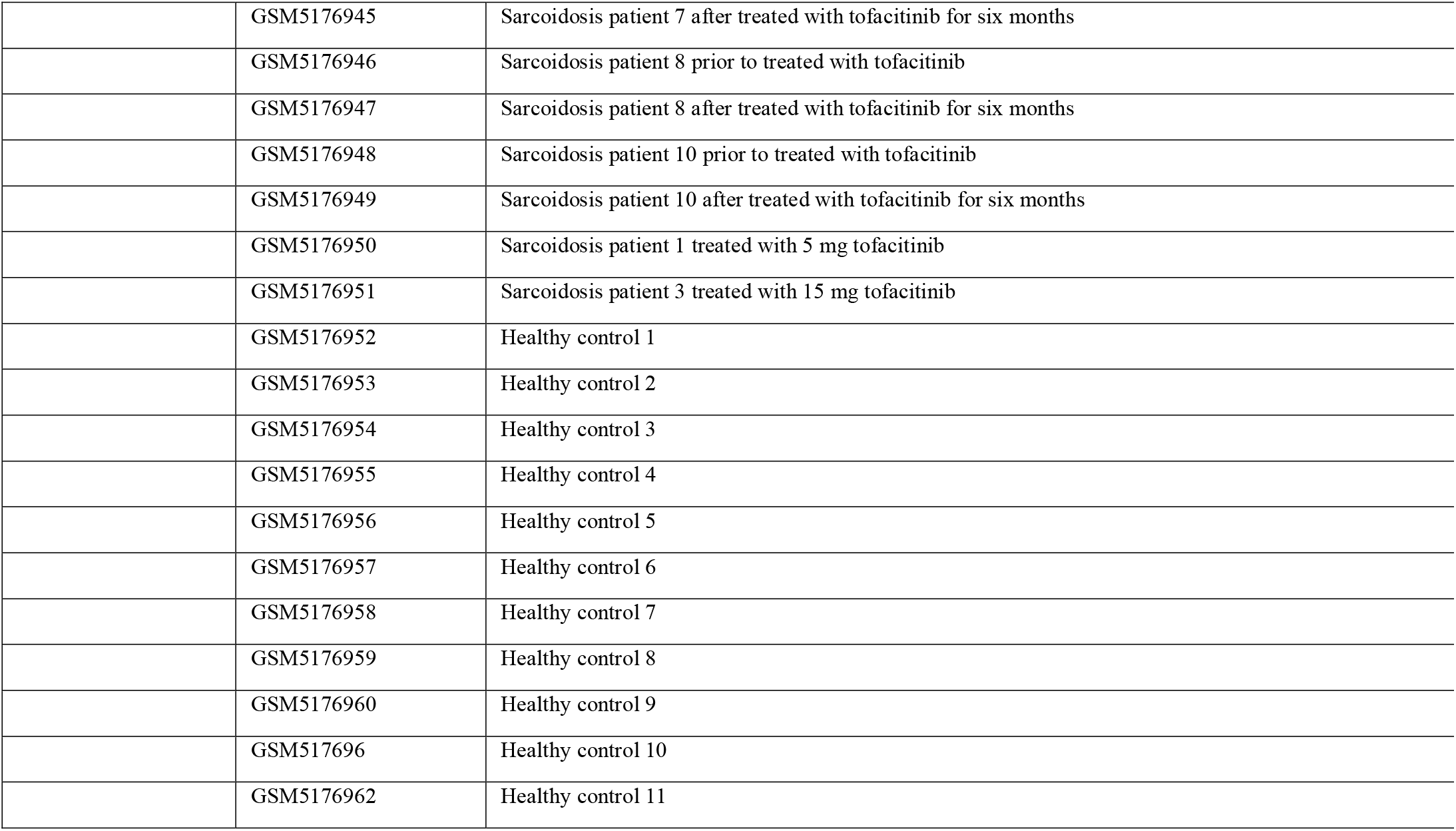
Samples available on Gene Expression Omnibus.

## SUPPLEMENTARY FIGURES

**Figure S1.**
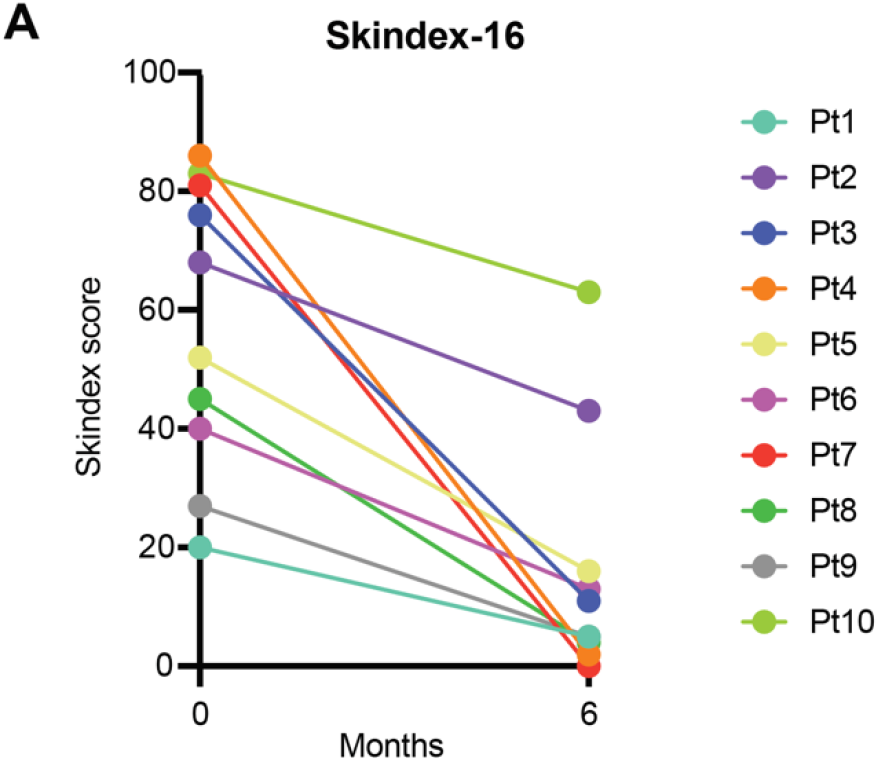
Changes in skin-related quality of life metric (Skindex-16). The Skindex-16 metric was administered at baseline and again after 6 months of tofacitinib.

**Figure S2.**
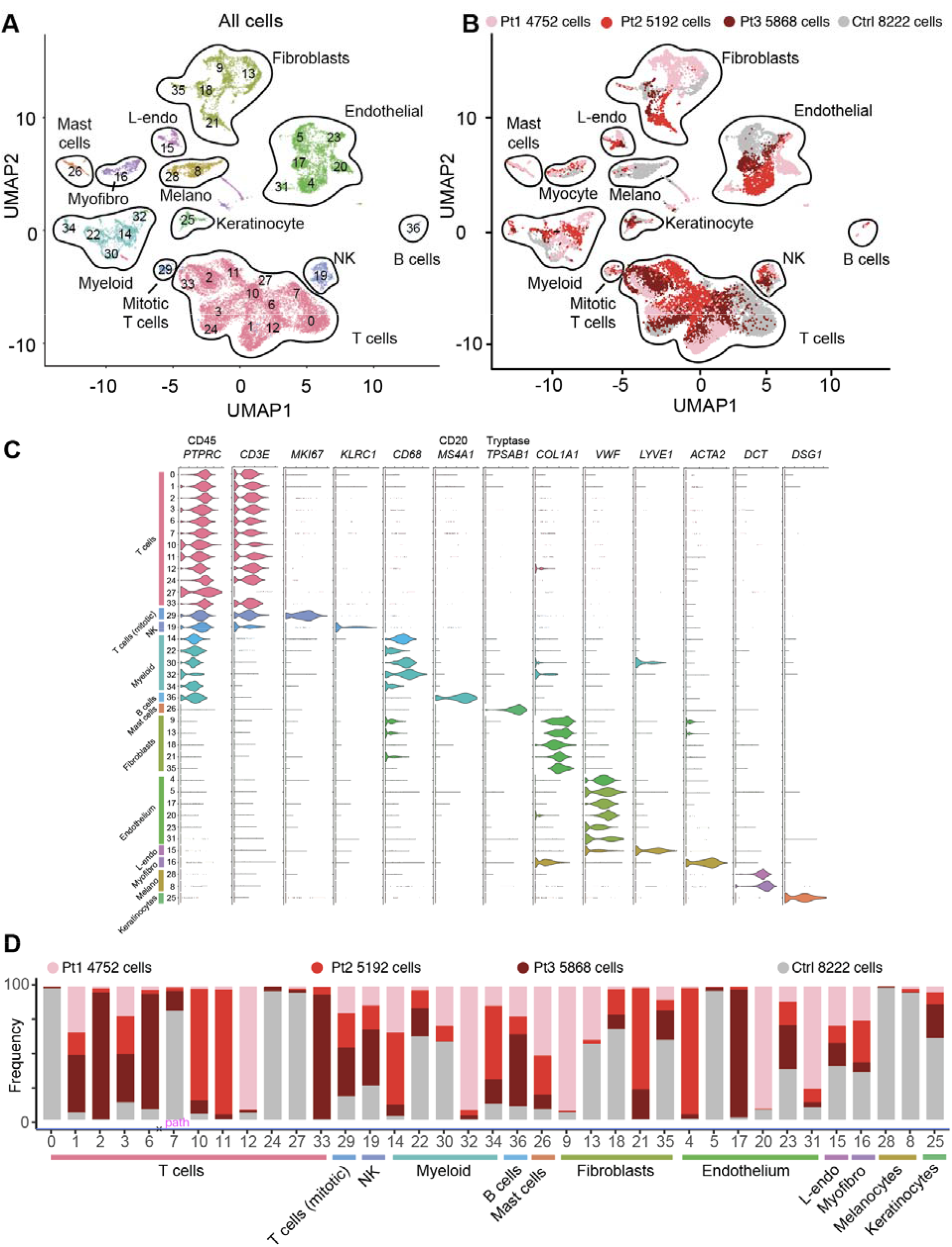
Summary of all cells analyzed using scRNA-seq in sarcoidosis and control skin. **A**. UMAP projection of scRNA-seq data showing clustering of all cells, colored by cell type. **B**. UMAP projection of scRNA-seq data in **A**, colored by condition/library. **C**. Violin plots showing cell-lineage markers used to classify cell types in **A** and **B. D**. Histograms showing contribution of each library/condition to each cluster. NK: natural killer cell, L-endo: lymphatic endothelium, Myofibro: myofibroblast.

**Figure S3.**
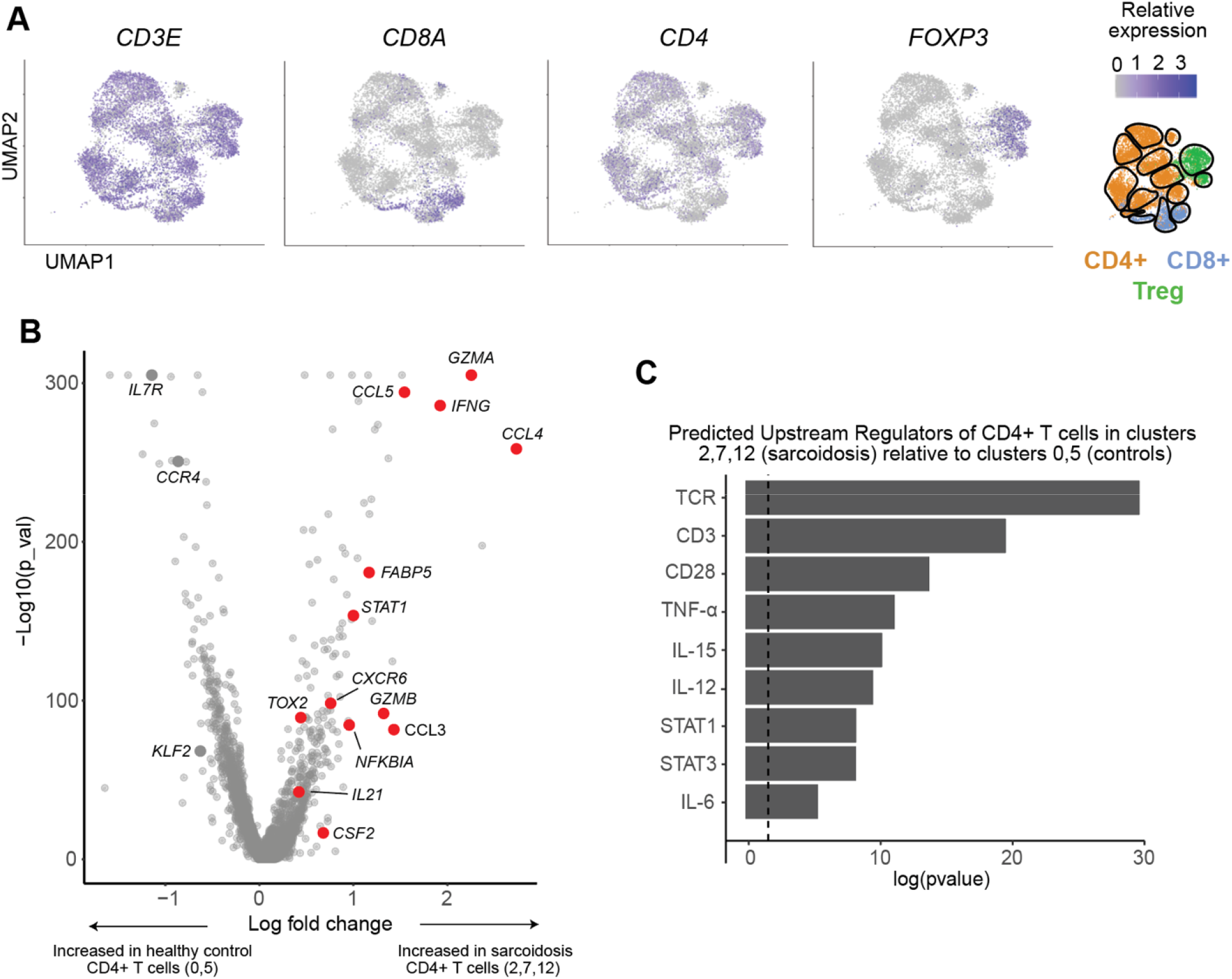
Analysis of T cells in scRNA-seq experiments. **A**. UMAP projections of T cell clusters from scRNA-seq experiments (corresponding to Figure 3A) showing relative expression levels of selected genes. **B**. Volcano plot showing the most differentially expressed genes between CD4+ T cells in sarcoidosis (clusters 2,7,12) versus CD4+ T cells in controls (clusters 0,5), corresponding to Figure 3A. **C**. Histogram showing selected predicted upstream regulators of CD4+ T cells in sarcoidosis (clusters 2,7,12) versus CD4+ T cells in controls (clusters 0,5) as determined by IPA. Significance cutoff of p<0.001 is shown by a dotted vertical line.

**Figure S4.**
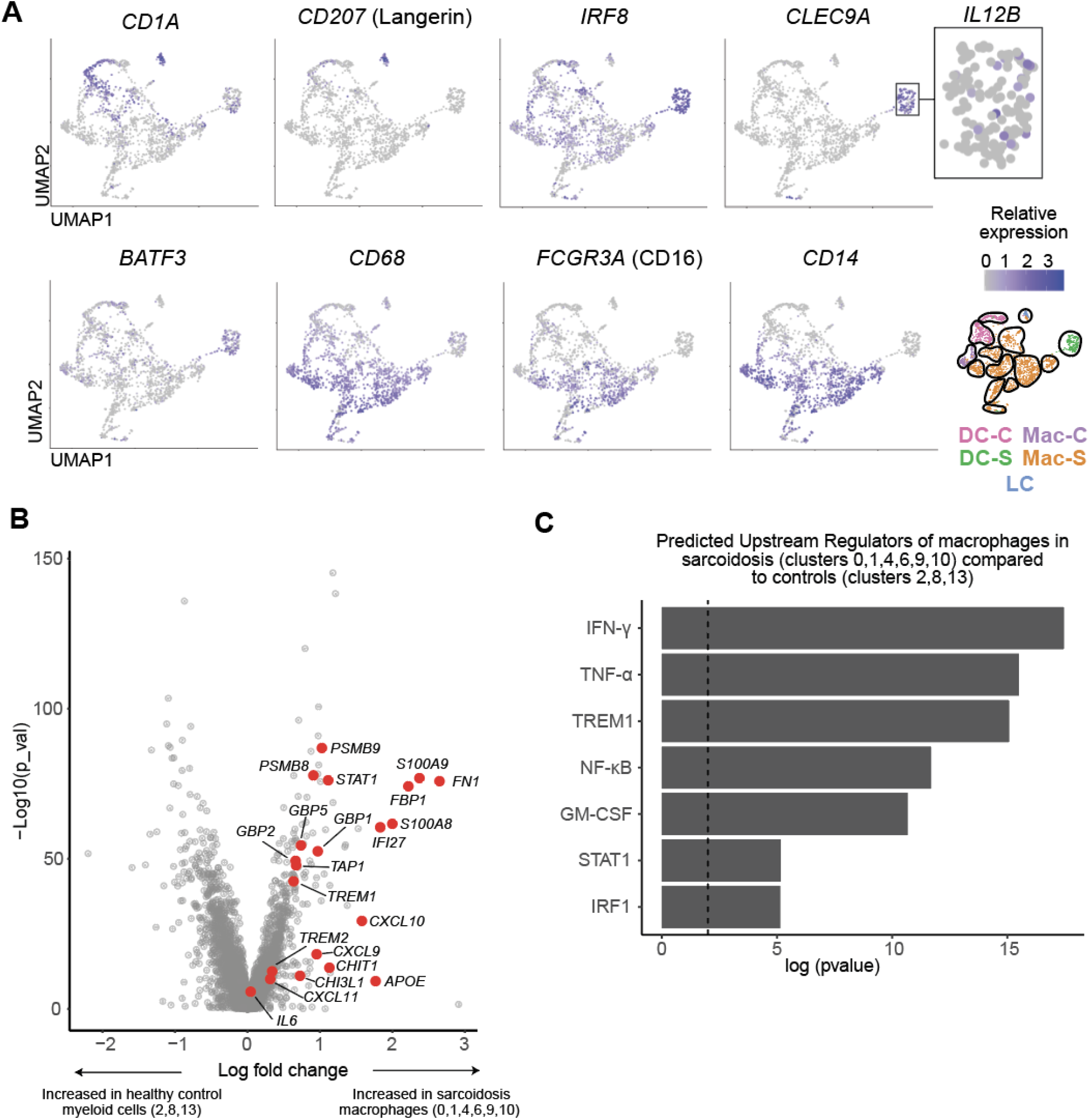
Analysis of myeloid cells in scRNA-seq experiments. **A**. UMAP projections of myeloid cell clusters from scRNA-seq experiments (corresponding to Figure 3B) showing relative expression of selected genes. Inset shows *IL12B* expression in cluster DC-S (DCs with a cDC1 phenotype in sarcoidosis). **B**. Volcano plot showing the most differentially expressed genes between macrophages in sarcoidosis (clusters 0,1,3,4,6,7,9,10,11) versus myeloid cells in controls (clusters 2,8,13). **C**. Histogram showing selected predicted upstream regulators in macrophages in sarcoidosis (clusters 0,1,3,4,6,7,9,10,11) versus myeloid cells in controls (clusters 2,8,13) as determined by IPA. Significance cutoff of p<0.001 is shown by a dotted vertical line.

**Figure S5.**
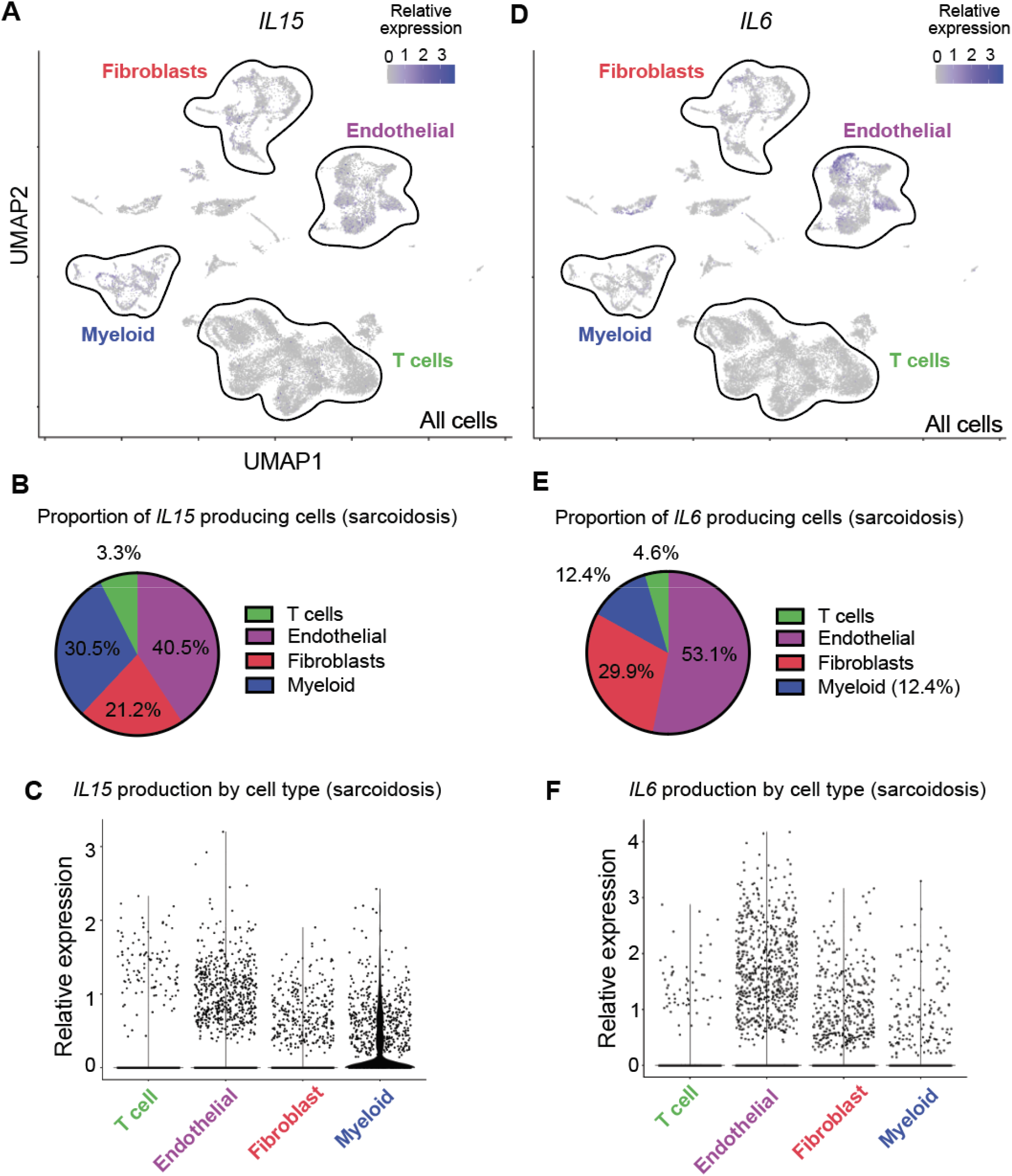
Analysis of all cells in scRNA-seq experiments. **A**. UMAP projection of all cells from scRNA-seq experiments (corresponding to Figure S2A,B) showing relative expression of *IL15*. **B**. Pie chart showing relative proportions of *IL15* producing cells in sarcoidosis libraries. **C**. Violin plot showing relative expression of IL15 by cell type in sarcoidosis libraries. **D**. UMAP projection of all cells from scRNA-seq experiments (corresponding to Figure S2A,B) showing relative expression of *IL6*. **E**. Pie chart showing relative proportions of *IL6* producing cells in sarcoidosis libraries. **F**. Violin plot showing relative expression of IL6 by cell type in sarcoidosis libraries.

**Figure S6.**
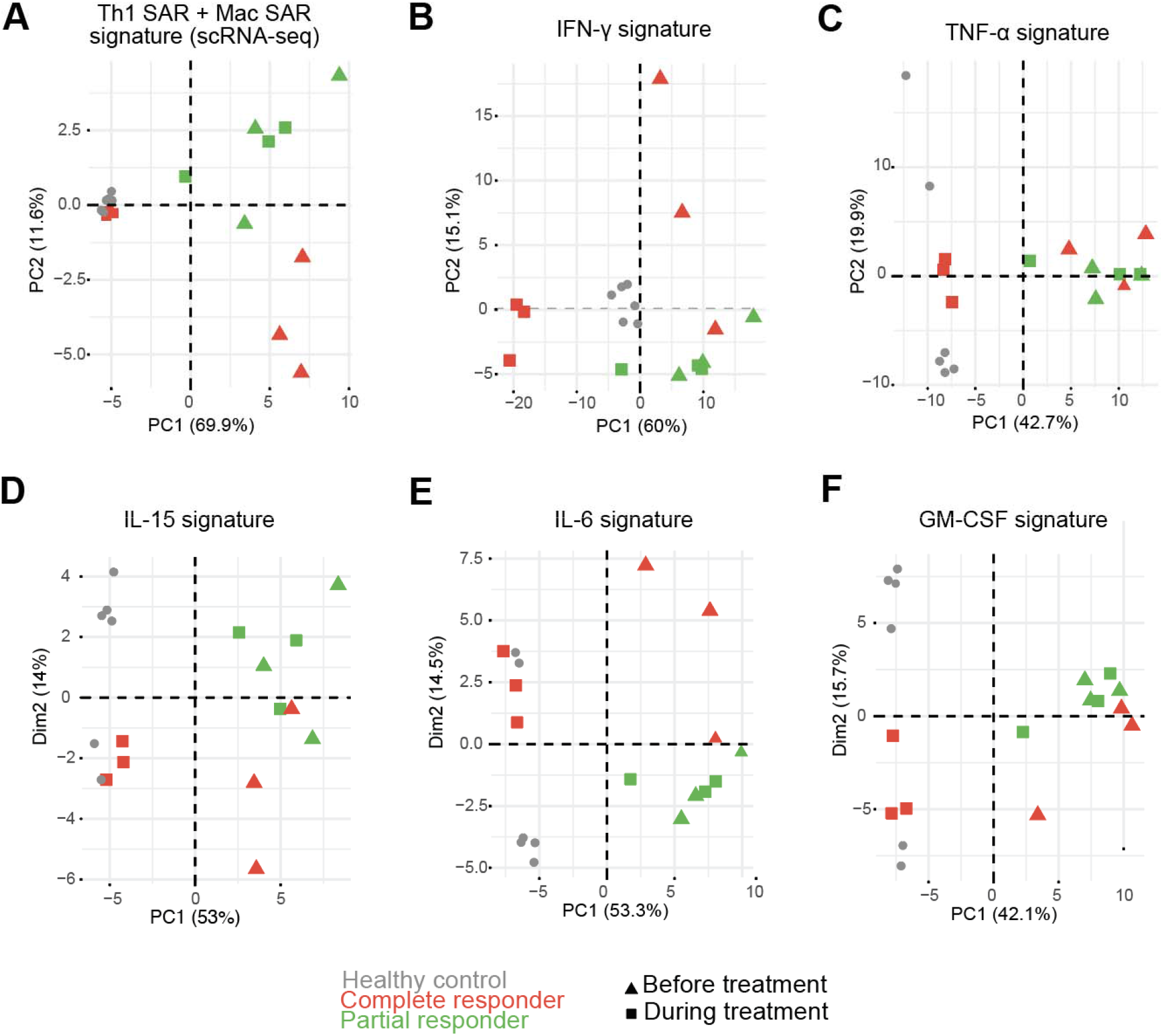
Principal component analysis of gene expression data with various gene sets. Analysis of gene expression data from skin in complete responders (CR) and partial responders (PR) relative to healthy controls. **A**. Macrophage (Mac-SAR) and T cell (Th1 SAR) activation signature genes from scRNA-seq analysis (Figure 3C) were used to perform principal component analysis. **B-F** Cytokine response signature gene sets (Figure 3J) were used to perform principal component analysis.

